# Impact of Plasma pTau181 Levels on Clinician Diagnostic Confidence and Management in Memory and Cognition Clinics: A Multi-site Before-and-After Study

**DOI:** 10.1101/2025.05.06.25327054

**Authors:** Johannes C. Michaelian, Azadeh Feizpour, James C. Vickers, Jane E. Alty, Jessica M. Collins, Vincent Doré, Catriona Ireland, Anna E. King, Ralph N. Martins, Michael Woodward, Sharon L. Naismith, Christopher C. Rowe

## Abstract

**INTRODUCTION:** Recent advances allow blood tests to detect key proteins linked to Alzheimer’s disease (AD).

**METHODS:** In this before-and-after study across three Australian Memory and Cognition Clinics, we evaluated the impact on clinicians’ diagnostic confidence and management following disclosure of routine patients’ AD probability, using predefined plasma pTau181 thresholds set at 90% sensitivity and 90% specificity for amyloid-β (Aβ) PET positivity.

**RESULTS:** 113 participants (age:71.2±8.4; MMSE:27.7±2.5) with dementia (n=17, 15.0%), mild cognitive impairment (n=48, 42.5%) and subjective cognitive decline (n=48, 42.5%) were enrolled. Blood test results were ‘probably negative’, n=81, 71.7%; indeterminate, n=24, 21.2%; ‘probably positive’, n=8, 7.1%. In 12 cases (10.6%), pTau181 changed clinician diagnosis and increased mean diagnostic confidence from low-to-moderate (61%) to moderate-to-high (80%). Aβ-PET in 40 participants showed plasma pTau181 improved diagnostic accuracy by 5%.

**DISCUSSION:** This study demonstrates the benefits of plasma pTau181 in real-world clinical practice particularly when diagnostic confidence is only low-to-moderate.

## BACKGROUND

The accurate and timely diagnosis of Alzheimer’s disease (AD), and particularly the early prodromal stage referred to as mild cognitive impairment (MCI), remains a significant challenge [1]. While multidisciplinary specialist assessment services for dementia and cognitive decline, often referred to as Memory and Cognition Clinics, are best placed to provide comprehensive assessment, access and availability to these services are limited [2–4]. Adding to this, approximately 50% of dementia cases go undetected in primary care [5, 6], with average delays of more than three years between initial symptom presentation and formal diagnosis [5, 6]. Moreover, while cognitive and clinical assessments are the foundation for diagnosis, they lack sensitivity in determining the underlying mechanisms of disease, particularly in prodromal stages [7].

In this regard, while the recent advancements in biomarker-based approaches, including positron emission tomography (PET) imaging and cerebrospinal fluid (CSF) analysis have facilitated the investigation and quantification of the neuropathological hallmarks of disease *pre-mortem*, namely amyloid-β (Aβ) and phosphorylated-tau, these investigations are invasive, costly, and not easily accessible in routine clinical practice [8, 9]. It is here that recent advancements in blood-based biomarkers have emerged as a promising tool for detecting underlying AD neuropathology [10]. Indeed, plasma biomarkers targeting phosphorylated tau (pTau), have demonstrated strong concordance with *in-vivo* gold standard Aβ-PET and CSF assays [11]. Plasma pTau181 has been shown to accurately predict Aβ-PET positivity with high sensitivity and specificity in a population of older adults undergoing routine assessment [12, 13], supporting it as a potential biomarker to improve the diagnostic process. However, despite these advancements, there remains limited real-world studies evaluating the impact of disclosing routine patient’s AD probability, determined by plasma pTau181 levels, on clinicians’ diagnostic confidence and management.

Therefore, in this before-and-after study across three Australian Memory and Cognition Clinics, we aimed to evaluate the impact of disclosing a routine patient’s AD probability, determined by plasma pTau181 levels on clinician:

1. Diagnosis and diagnostic confidence;
2. Management decisions and management confidence; and
3. Perspectives on the enablers and barriers in implementing a blood test for AD

## METHODS

### Study Design and eligibility criteria

Patients attending a Memory and Cognition Clinic (n = 3; Austin Health Cognitive Dementia and Memory Service [CDAMS] in Melbourne, Healthy Brain Ageing [HBA] Clinic in Sydney, and ISLAND Clinic in Hobart) for routine assessment were consecutively enrolled in this multi-centre, before-and-after study. To facilitate a ‘real-world’ study, participants were included if they were attending the Memory and Cognition Clinic for an assessment for cognitive concerns or dementia, aged 50 years or older, presented with a Mini-Mental State Examination (MMSE) score 23 or greater, and were able to identify a ‘support person’ (e.g. family member, spouse, caregiver) to be involved in the study. Following ascertainment of eligibility and informed consent, participants provided a fasting blood-sample and completed brief self-report questionnaires to ascertain current mental health well-being (WHO-5 Well-Being Index, [14]), quality of life (EQ-5D-5L, [15]), depressive symptoms (15-item Geriatric Depression Scale, [16]) and overall disability (WHO Disability Assessment Schedule 2.0, [17]). Where participants had previously provided a fasting blood-sample or completed the same questionnaires as part of their routine assessment at the Memory and Cognition Clinic, consent was sought to utilise already collected data, in addition to brief clinical and demographic history relevant to the outcomes of this study.

#### Participant diagnosis and management pre- and post-blood test disclosure

During patient case conference at the Memory and Cognition Clinic, and before disclosing blood test results, clinicians completed a structured questionnaire to document their diagnosis, management plan, and level of confidence (see **Supplementary Text 1**). For both HBA and ISLAND clinics, as patient case-conference was conducted on the day of assessment, blood test results were not available to be disclosed to study clinicians until approximately 4-6 weeks after; however as Austin Health CDAMS conducted case-conference approximately 12 weeks following patient assessment to allow for the collection and reporting of further investigations (i.e., MRI and FDG-PET imaging) – clinician diagnosis and management, including levels of confidence were captured immediately prior to disclosing blood test results. Where Aβ-PET imaging results were available for participants enrolled in the study, the Aβ-PET results were withheld until following blood test result disclosure and impact assessment. For all sites, following blood test disclosure (see **Figure 1** for blood test result examples), the same study clinician completed a structured questionnaire (see **Supplementary Text 2**) to evaluate whether the blood test result had an impact on their patient’s diagnosis, their management plan, including their level of confidence.

**FIGURE 1.**
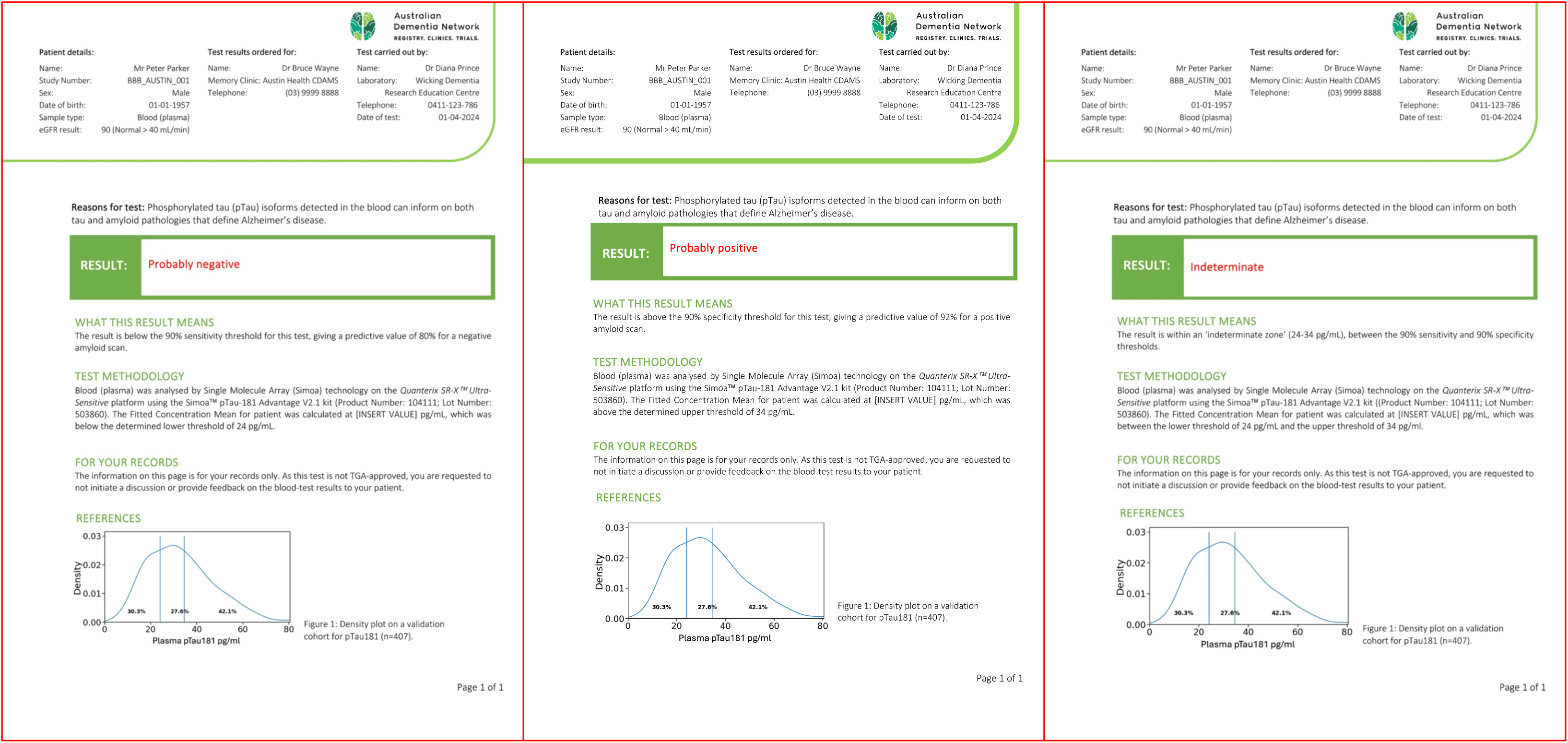
Examples of blood test reports provided to study clinicians presenting plasma ptau181 results and their corresponding interpretations.

#### Study clinician close out survey

At the conclusion of the study, all study clinicians were provided an opportunity to complete a close-out survey to capture the enablers and barriers toward the clinical implementation for a blood-test for dementia (see **Supplementary Text 3**).

### Study approval and registration

The study ‘Implementing Blood-Based Biomarkers into Memory Clinics: A Before and After Study Evaluating the Impact of Blood-Based Biomarkers on Clinician Diagnostic Confidence and Management’ was approved by the Human Ethics Review Committee (RPAH Zone) of the Sydney Local Health District (approval/protocol number: X21-0427). The study was prospectively registered on the Australian and New Zealand Clinical Trial Registry (ACTRN12622000515796, 31 March 2022). All participants provided written informed consent to take part in the study, and all data was obtained in compliance with the Helsinki Declaration.

### Plasma pTau181 threshold development and analysis

Prior to study commencement, a validation study was performed on a combined AIBL and ADNeT cohort (n = 407) to determine two thresholds of plasma pTau181 concentration set at 90% sensitivity and 90% specificity for Aβ-PET positivity (≥35 Centiloids [CL]). The 90% sensitivity threshold corresponded to 24 pg/ml and the 90% specificity threshold corresponded to 34 pg/ml. Underlying disease prevalence (% PET+) for the entire validation cohort was 64%. Based on the two-threshold approach, 30% of participants fell in the low range, 28% in the indeterminate range, and 42% in the high range. Positive percent agreement was 92% and negative percent agreement was 80%. Based on these criteria, and as illustrated in Figure 1, one of three possible results were communicated to clinicians:

- Probably positive (> 34 pg/mL): the result is above the 90% specificity threshold for this test, giving a predictive value of 92% for a positive Aβ-PET scan (CL value ≥ 35);
- Probably negative (< 24 pg/mL): the result is below the 90% sensitivity threshold for this test, giving a predictive value of 80% for a negative Aβ-PET scan (CL value < 35);
- Indeterminant (24-34 pg/mL): the result is within an ‘indeterminate zone’ (24-34 pg/mL), between the 90% sensitivity and 90% specificity thresholds.

For both the validation study and following study commencement, plasma was extracted from blood collected in an EDTA vacuum tube, stored at -80°C, and thawed before use. Utilising Single Molecule Array (Simoa) technology on the Quanterix SR-X^TM^ Ultra-Sensitive platform, pTau181 was analysed using the Simoa^TM^ pTau-181 Advantage V2.1 kit, according to manufacturer instructions and recommendations (i.e., all samples were run in duplicate). Duplicate results were used if the coefficient of variation (CV%) was <20%; otherwise, plasma samples were reanalysed until a CV% <20% was achieved.

### Amyloid-β PET imaging and brain amyloid quantification for subsample of participants

All participants at one site (Austin Health, Melbourne) also underwent an Aβ-PET scan. This substudy was approved by the Austin Health Human Research Ethics Committee (HREC approval/protocol number: HREC/89415/Austin-2023) and required participants to review and sign an additional consent form specifically for the scan. A 20-minute Aβ-PET scan was performed 50 minutes after injecting 200 MBq of ^18^F-NAV4694 produced in the radiochemistry laboratory at Austin Health. Using CapAIBL [18], Aβ-PET scans were spatially normalized, and the standard Centiloid (CL) method was applied [19, 20], with the whole cerebellum mask as the reference region. A CL value of 35 was chosen to identify a positive Aβ scan as used for the threshold development.

### Statistical analyses

Descriptive analyses (i.e., frequencies, means, standard deviations, correlations) were performed in SPSS Version 29.0 (SPSS Inc., Chicago, IL, USA) to provide an overview of the data. Scatter plots were generated in R Statistical Software (R Version 4.4.2; R Core Team 2024), including concordance analyses which were conducted using McNemar’s test and proportion comparison.

## RESULTS

### Baseline Participant and Study Site Characteristics

As illustrated in **Figure 2**, participant recruitment across three study sites (Austin Health CDAMS, HBA Clinic and ISLAND Clinic) occurred between February and October 2024, with a total of 113 participants enrolled in the study. Moreover, as reported in **Table 1**, two study sites were considered ‘hybrid’ (i.e., clinics embedded within universities, funded via grants and Medicare) receiving the majority (>90%) of patient referrals from General Practitioners (GPs), while one was located in a metropolitan public hospital and received patient referrals from both GPs (45%) and medical specialists (55%). Among the eight clinicians who participated in the study, the majority were geriatricians (n = 7, 87.5%).

**FIGURE 2.**
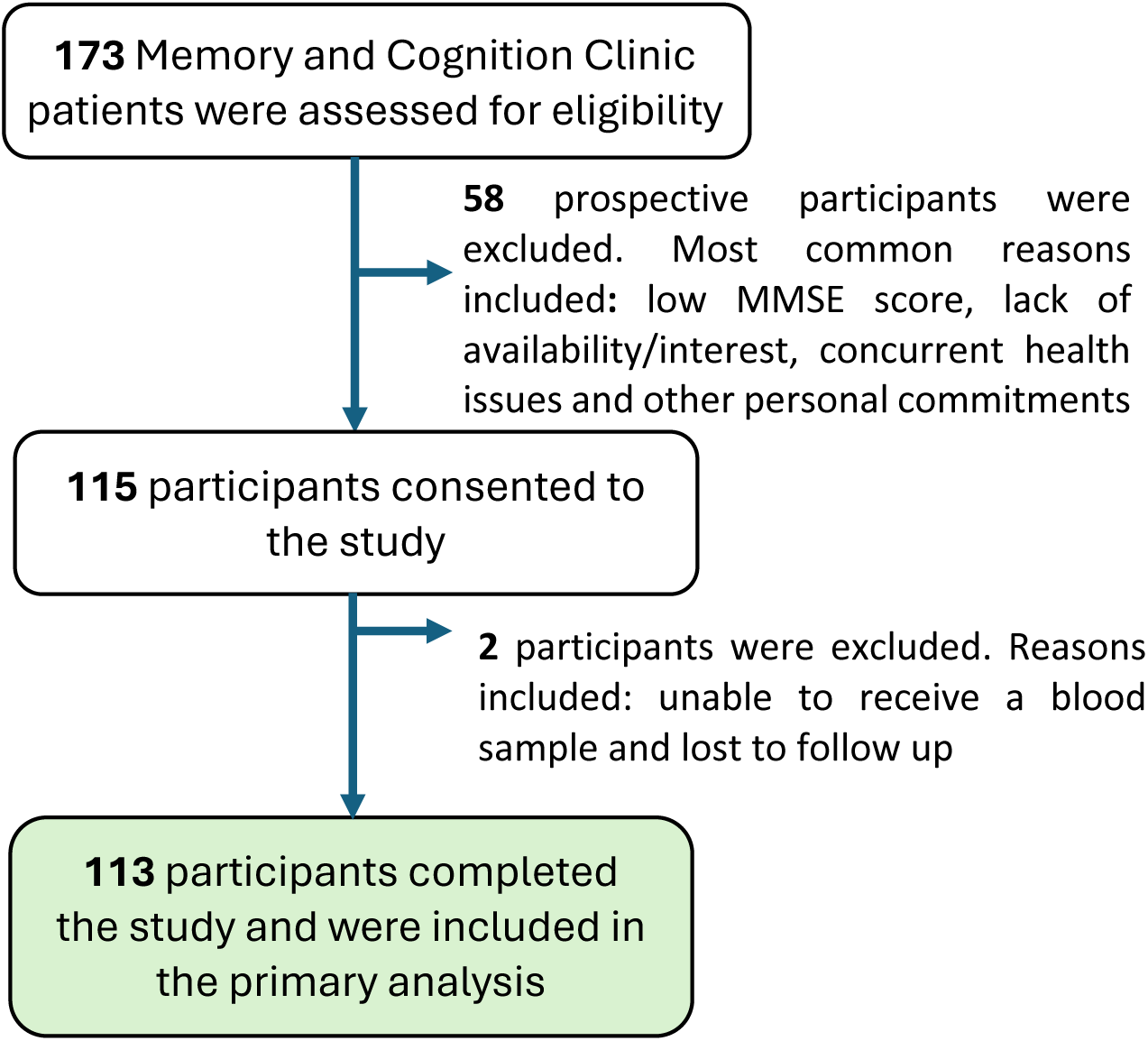
Participant recruitment and enrolment flowchart between February and October 2024.

**TABLE 1.** Characteristics of participating clinics

| **Characteristic** | **Austin Health**<br>**CDAMS** | **HBA Clinic** | **ISLAND Clinic** |
| --- | --- | --- | --- |
| **Participating clinicians, n** | 4 | 1 | 3 |
| **Participants enrolled, n** | 40 | 40 | 33 |
| **Clinician specialty, n (%)** |  |  |  |
| Geriatrician | 3 (75.0) | 1 (100.0) | 3 (100.0) |
| Neurologist | 1 (25.0) | 0 (0) | 0 (0) |
| **Clinic type** | Public | Hybrid | Hybrid |
| **Clinic locality**<br>**(ABS classification)** | Metropolitan | Metropolitan | Regional |
| **Referral source,**<br>**n (%)** |  |  |  |
| General Practitioner | 18 (45.0) | 40 (100.0) | 31 (93.9) |
| Other specialist | 22 (55.0) | 0 (0) | 2 (6.1) |

*Note.* Hybrid clinics are embedded within universities, funded via grants with additional use of Medicare.
Abbreviations: CDAMS, Cognitive Dementia and Memory Service; HBA, Healthy Brain
Ageing; ABS, Australian Bureau of Statistics

As detailed in **Table 2**, participants across all study sites were on average 71 years old, with participants enrolled at the HBA Clinic significantly younger (mean age = 67.5 years) and more likely to be currently working (n = 20, 50.0%). Compared to Austin Health CDAMS, a significantly higher proportion of female participants were enrolled at HBA (n = 31, 77.5%) and ISLAND Clinics (n = 21, 63.6%). While clinical diagnoses and classifications varied across sites, the ISLAND Clinic had a significantly higher proportion of dementia cases (n = 9, 27.3%); whereas the HBA Clinic had significantly fewer participants with dementia (n = 2, 5.0%) and a higher proportion with subjective cognitive decline (SCD, n = 32, 80.0%). MMSE scores were consistent across sites (mean = 27.7), as were education levels (mean = 13.7 years) and the proportion of participants who spoke English at home (mean = 90.2%). Notably, nearly two-thirds of participants (n = 73, 65.2%) were born in Australia. While estimated glomerular filtration rate (eGFR) aligned with the average age of participants across study sites (mean = 76.7) [21], body mass index (BMI) values (mean = 28.4) would be considered overweight/pre-obese according to current WHO general classification criteria [22]. Furthermore, according to the Index of Relative Socio-Economic Advantage and Disadvantage (IRSAD) calculated for all participants [23], ISLAND participants had significantly lower socio-economic conditions (mean = 4.3) when compared to participants enrolled at the HBA Clinic (mean = 9.0) and Austin Health CDAMS (mean = 7.6).

**TABLE 2.**
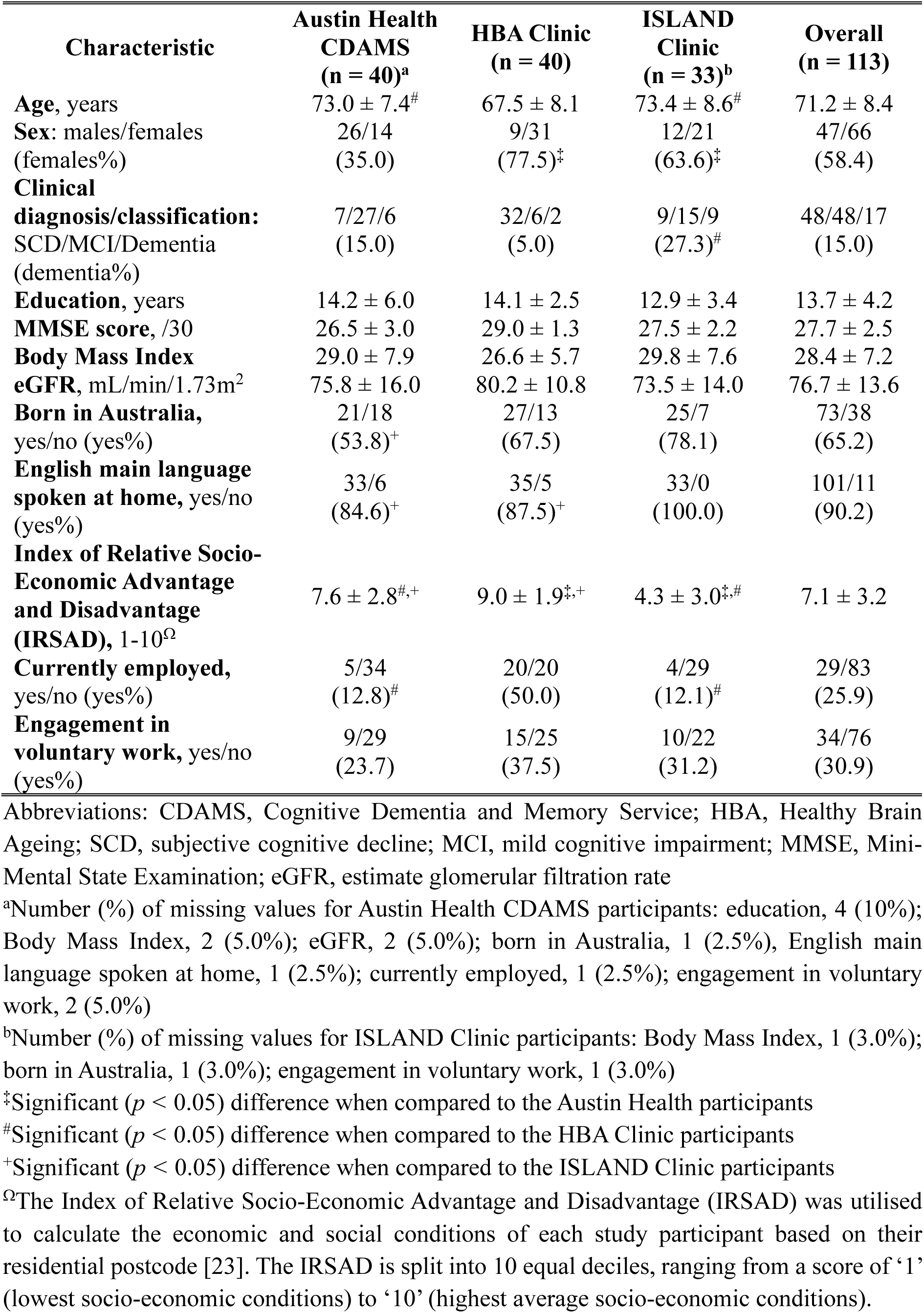
Baseline characteristics of study participants across clinics (mean ± SD)

**Table 3** presents participant self-reported quality of life, mental and physical health data across study sites. Overall, mental health well-being (WHO-5, 15.1 ± 5.7) and symptoms of depression (GDS-15, 6.0 ± 2.7) were normal, with no significant differences between study site populations. While self-reported overall health on the day of assessment was relatively high across all sites (EQ-5D-5L, 74.1 ± 19.5), mild disability in everyday functioning was reported (WHODAS 2.0, 16.9 ± 14.0), a finding consistent across all sites. Across EQ-5D-5L subdomains, Austin Health CDAMS participants reported significantly greater problems in mobility, self-care, and usual activities compared to HBA and ISLAND Clinic participants. Additionally, both Austin Health CDAMS and ISLAND Clinic participants reported greater problems around pain/discomfort and anxiety/depression than HBA Clinic participants.

**TABLE 3.**
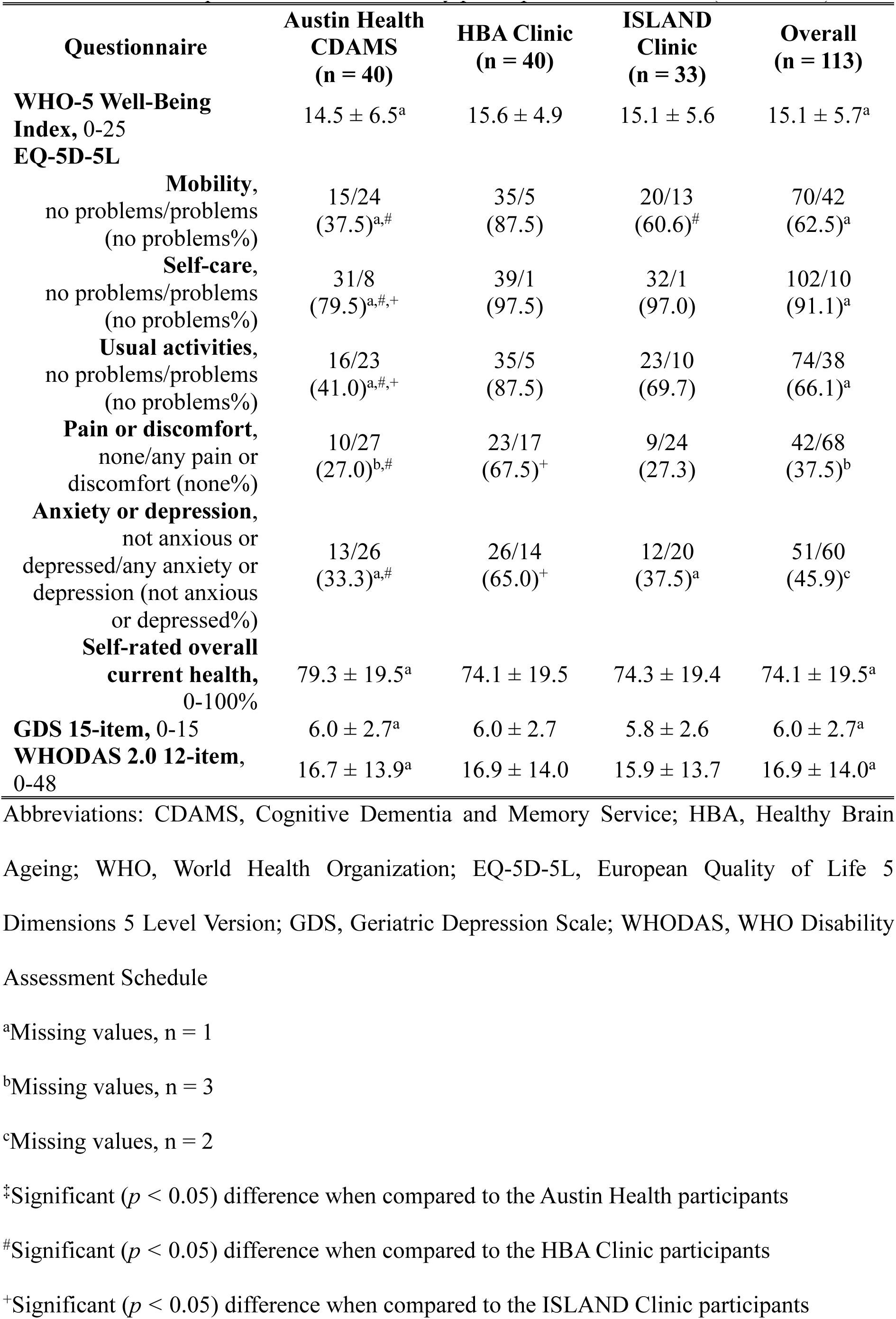
Baseline questionnaire data of study participants across clinics (mean ± SD)

### Plasma pTau181 levels across clinical classifications

Comparing initial clinical classifications/diagnoses across all study sites, plasma pTau181 levels were significantly lower in participants classified with SCD (mean ± SD, 16.6 ± 15.6 pg/mL) compared to participants with MCI (22.7 ± 20.5 pg/mL, *t*(73.8) = -3.52, *p* < 0.001) and dementia (24.6 ± 23.5 pg/mL, *t*(20.9) = -3.39, *p* = 0.003). Plasma pTau181 levels did not significantly differ between participants classified with MCI (22.7 ± 20.5 pg/mL) and diagnosed with dementia (24.6 ± 23.5 pg/mL, *t*(63) = -0.67, *p* = 0.504).

### Changes in diagnostic and management confidence pre- and post-blood test disclosure

Of the 113 participants enrolled in the study, and based on the validation study, most blood test results received were ‘probably negative’ (n = 81, 71.7%: SCD, n = 43 [53.1%]; MCI, n = 29 [35.8%]; dementia, n = 9 [11.1%]), followed by indeterminate (n = 24, 21.2%: SCD, n = 5 [20.8%]; MCI, n = 14 [58.4%]; dementia, n = 5 [20.8%]) and ‘probably positive’ (n=8, 7.1%: SCD, n = 0 [0%]; MCI, n = 5 [62.5%]; dementia, n = 3 [37.5%]). As shown in **Table 4**, mean diagnostic and management confidence following the initial clinical assessment was moderate-to-high across all sites, at 77.2% and 81.1%, respectively. In the sample where clinicians reported diagnostic change following disclosure of blood test results (n = 12, 10.6%: ‘probably negative’, n = 7; ‘probably positive,’ n = 2; indeterminate, n = 3), diagnostic confidence increased in these patients from on average low-to-moderate (60.8%) to on average moderate-to-high (79.6%), a mean increase of 18.8%. Similarly, in the sample where clinicians reported management change following disclosure of blood test results (n = 8), their management confidence increased in these patients from low-to-moderate (66.9%) to moderate-to-high (83.9%), a mean increase of 17.0%. In approximately a quarter of participants (n = 30, 26.5%), the blood test results did not change the clinician’s diagnosis or management but increased overall mean confidence, with diagnostic confidence rising from 69.0% to 81.5% and management confidence from 72.6% to 80.7% (**Table 5**).

**TABLE 4.** Changes in diagnostic and management confidence pre- and post-blood test disclosure: for patients *with* post-blood test diagnosis or management changes (mean ± SD)

| Clinic | Diagnostic confidence (%) |  |  | Management confidence (%) |  |  |
| --- | --- | --- | --- | --- | --- | --- |
|  | Entire sample *pre*-blood test (n = 113) | Sample reporting change following disclosure *pre*-blood test (n = 12) | Sample reporting change following disclosure *post*-blood test (n = 12) | Entire sample *Pre*-blood test (n = 113) | Sample reporting change following disclosure *pre*-blood test (n = 8) | Sample reporting change following disclosure *post*-blood test (n = 8) |
| **Overall** | 77.2 ± 17.6 | 60.8 ± 18.6 | 79.6 ± 15.1 | 81.1 ± 15.2 | 66.9 ± 21.7 | 83.9 ± 12.2 |
| **Austin Health CDAMS** | 64.3 ± 16.6 | 57.4 ± 16.6<sup>a</sup> | 77.5 ± 14.5<sup>a</sup> | 68.2 ± 15.6 | 57.5 ± 15.4<sup>c</sup> | 79.3 ± 10.3<sup>c</sup> |
| **HBA Clinic** | 91.1 ± 6.7 | 77.5 ± 11.9<sup>b</sup> | 90.0 ± 10.4<sup>b</sup> | 93.6 ± 3.0 | 95.0 ± 0<sup>d</sup> | 97.5 ± 3.5<sup>d</sup> |
| **ISLAND Clinic** | 76.1 ± 15.4 | - | - | 81.5 ± 9.7 | - | - |

*Note.* Change in diagnostic confidence post-blood test disclosure included both an increase (n = 9; mean: 20%) and decrease (n = 3; mean: 15%) in confidence. There were no decreases in management confidence following post-blood test disclosure (n = 8).
Abbreviations: CDAMS, Cognitive Dementia and Memory Service; HBA, Healthy Brain Ageing
<sup>a</sup>Austin Health CDAMS, n = 10; <sup>b</sup>HBA Clinic, n = 2; <sup>c</sup>Austin Health CDAMS, n = 6; <sup>d</sup>HBA Clinic, n = 2

**TABLE 5.** Changes in diagnostic and management confidence pre- and post-blood test disclosure: for patients *without* post-blood test diagnosis or management changes (mean ± SD)

| Clinic | Diagnostic confidence (%) |  |  | Management confidence (%) |  |  |
| --- | --- | --- | --- | --- | --- | --- |
|  | Entire sample *pre*-blood test (n = 113) | Sample reporting No change following disclosure *pre*-blood test (n = 30) | Sample reporting No change following disclosure *post*-blood test (n = 30) | Entire sample *pre*-blood test (n = 113) | Sample reporting No change following disclosure *pre*-blood test (n = 20) | Sample reporting No change following disclosure *post*-blood test (n = 20) |
| **Overall** | 77.2 ± 17.6 | 69.0 ± 17.6 | 81.5 ± 19.3 | 81.1 ± 15.2 | 72.6 ± 13.0 | 80.7 ± 15.4 |
| **Austin Health CDAMS** | 64.3 ± 16.6 | 68.2 ± 16.6<sup>a</sup> | 86.0 ± 11.8<sup>a</sup> | 68.2 ± 15.6 | 72.8 ± 13.3<sup>d</sup> | 81.2 ± 15.7<sup>d</sup> |
| **HBA Clinic** | 91.1 ± 6.7 | 83.8 ± 5.3<sup>b</sup> | 82.5 ± 16.6<sup>b</sup> | 93.6 ± 3.0 | -<sup>e</sup> | -<sup>e</sup> |
| **ISLAND Clinic** | 76.1 ± 15.4 | 50.0 ± 0 | 25.5 ± 0.7<sup>c</sup> | 81.5 ± 9.7 | 68<sup>f</sup> | 72<sup>f</sup> |

*Note.* Change in diagnostic confidence post-blood test disclosure included both an increase (n = 23; mean: 22.2%) and decrease (n = 7; mean: -19.6%) in confidence. Change in management confidence post-blood test disclosure included both an increase (n = 16; mean: 16.9%) and decrease (n = 4; mean: -27.3%) in confidence.
Abbreviations: CDAMS, Cognitive Dementia and Memory Service; HBA, Healthy Brain Ageing
<sup>a</sup>Austin Health CDAMS, n = 24; <sup>b</sup>HBA Clinic, n = 4; <sup>c</sup>ISLAND Clinic, n = 2; <sup>d</sup>Austin Health CDAMS, n = 19; <sup>e</sup>HBA Clinic, n = 0; <sup>f</sup>ISLAND Clinic, n = 1

### Amyloid-β PET imaging subsample enrolled at Austin Health CDAMS

All Austin Health CDAMS participants (n = 40) underwent Aβ-PET imaging and this revealed a low prevalence of AD pathology at 32.5% (**Figure 3**). Overall, in this subsample, concordance between Aβ-PET and a clinical diagnosis suggestive of an underlying AD process pre-blood test disclosure was assessed at 60% (n = 24), improving to 65% (n = 26) post-blood test disclosure. In the subset of participants where disclosure of blood test results changed clinician diagnosis (n = 10), pre-blood test diagnostic concordance with Aβ-PET was assessed at 30%, improving to 50% post-blood test disclosure.

**FIGURE 3.**
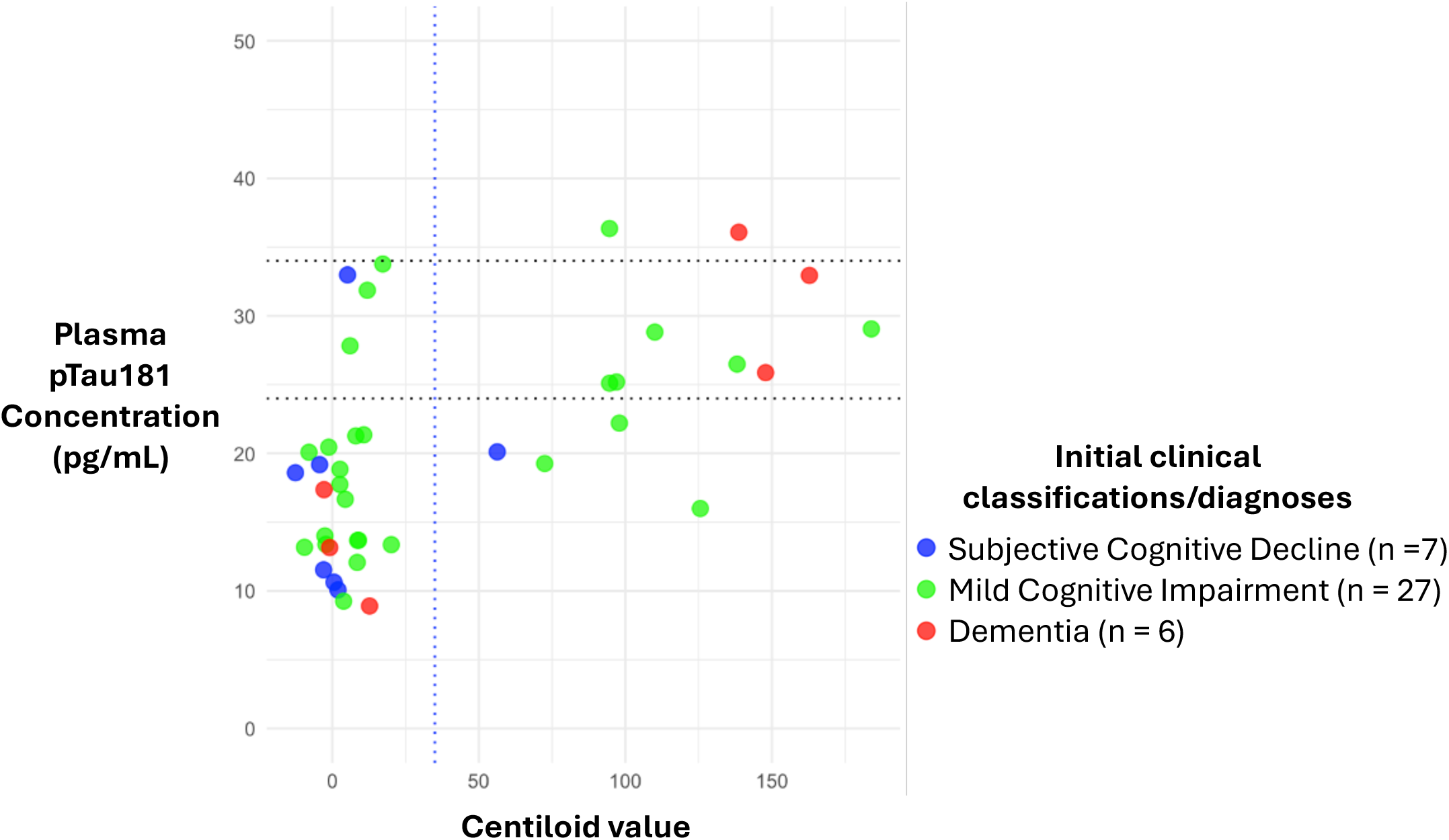
Scatter plot of brain amyloid-beta centiloid values against plasma pTau181 concentrations in a subset of participants (n = 40) who underwent amyloid-β PET scanning, separated by clinical classifications and diagnoses. Lower/upper horizontal cut-off: 24 and 34 pg/mL, respectively. Vertical cut-off: 35 Centiloids.

### Clinician perspectives on the enablers and barriers in implementing a blood test for AD

Following study close-out, two-thirds of clinicians (n = 6) completed a survey to ascertain the enablers and barriers towards the clinical implementation of a blood test for dementia. Overall experiences were mixed, with clinicians commenting that their experience was “excellent” and “interesting”, while others noted that it was “confusing” and “difficult to interpret” at times, for example:

- “I had an excellent experience with the blood-based biomarker information.”
- “I found it interesting but difficult to interpret in clinical context.”
- “Information on biomarkers is a welcome experience as we move towards clearer biological diagnosis for our patients. It was also interesting to integrate this information into the clinical diagnosis given our team has not used biomarkers previously.”
- “Confusing at times…”

Half of responding clinicians (n = 3) reported that blood test results provided greater confidence in their diagnosis, whereas half (n = 3) did not. Moreover, a third of responding clinicians (n = 2) reported that blood test results changed their management as a clinician. Two-thirds of clinicians (n = 4) commented that with further amendments they would recommend to other clinicians the routine use of a blood test for AD in clinical practice. Suggested amendments were mainly centred around the use of newer, more specific and sensitive blood-based biomarkers for an underlying AD process (e.g., pTau217). Barriers identified by clinicians were centred around “cost”, “availability/access” and “clinical correlation” and “limited understanding, for example:

- “Cost, availability”
- “Cost and ease of getting it done.”
- “Access to appropriate phlebotomy and pathology services in [location redacted]. I understand this is changing and it is available via [information redacted].”
- “Lack of clinical correlation”

Enablers identified by study clinicians mainly referenced the reporting of the blood test results, including:

- “Clear results and ranges, easy to see where the patient lay on the scale”
- “I found the printed results information sheet with interpretation of ranges educational and informative.”
- [it was] “quick, simple”

## DISCUSSSION

This real-world study provides novel insights into the clinical impact of disclosing AD probability based on plasma pTau181 levels for patients attending a Memory and Cognition Clinic. Our findings demonstrate that among patients who underwent a multi-disciplinary assessment and initially recorded a diagnosis with low-to-moderate confidence, disclosure of AD probability based on plasma pTau181 levels led to a change in underlying diagnosis for 10% of cases, increasing diagnostic confidence in these cases from low-to-moderate to moderate-to-high. Similarly, in 7% of cases disclosure of blood test results led to a change in management, increasing management confidence from low-to-moderate to moderate-to-high. Importantly, we report that two-thirds of study clinicians would recommend to their colleagues the clinical use of a blood test for AD in clinical practice with further amendment.

Consistent with previous reports, in this study, plasma pTau181 levels significantly increased across the dementia continuum, from SCD to MCI, as well as between participants with SCD and dementia [13, 24]. Although plasma pTau181 levels were, on average, higher in participants diagnosed with dementia compared to those with MCI, there was no significant difference likely due to the limited number of participants receiving a diagnosis of dementia enrolled across study sites (n = 17 dementia *versus* n = 48 MCI). Indeed, in a subsample of participants (n = 40) who underwent Aβ-PET imaging, the prevalence of Aβ-positivity (≥ 35 Centiloids) was low at 32.5%. This finding is in contrast to a meta-analysis of other cohort studies involving routine Memory and Cognition clinic patients, which reports an average Aβ-PET positivity rate exceeding 50% [25].

When taken together, the overarching diagnostic and management impact results of this study suggest a meaningful role for blood-based biomarkers in addressing clinical uncertainty, particularly for patients in the prodromal stage of disease, who may be indicated for an amyloid-targeting monoclonal antibody therapy for AD [26]. While our findings warrant further large-scale investigation across various Memory and Cognition Clinics, it may be suggested that the clinical implementation of a blood test for AD would enhance diagnostic accuracy in cases with diagnostic uncertainty (i.e., low-to-moderate confidence), and facilitate the triaging of eligible patients for a disease-modifying therapy.

Moreover, the novelty of this study lies in its real-world application of incorporating plasma pTau181 levels to determine AD probability for routine patients attending a Memory and Cognition Clinic for assessment. This approach addresses a key gap in the literature [27], and has provided valuable insights into clinician perspectives on the enablers and barriers to implementing a blood test for dementia in clinical practice. Examining these enablers and barriers, it was evident that the blood test report template (Figure 1) enabled the clear reporting and interpretation of results. However, clinician experiences varied - some found the report helpful, while others noted difficulties in interpretation or inconsistencies with patients’ clinical symptoms. Looking to the future, clinicians also raised concerns about the cost and accessibility of such a test in practice, as well as whether existing infrastructure (e.g., pathology collection services) could support its widespread implementation. Therefore, as the field moves ever closer to the clinical implementation of a diagnostic blood test for AD it is essential to ensure that reporting templates are standardised, and that clinicians have access to training and support to facilitate further diagnostic investigation with such blood tests.

This study was not without limitations. At the time of study design and commencement, investigators were constrained in their ability to measure plasma pTau217, which has been shown to demonstrate greater sensitivity, specificity and overall diagnostic accuracy for AD over plasma pTau181 [28]. Future studies should seek to replicate the current study’s methodology using pTau217 to better evaluate the impact of disclosing a routine patient’s AD probability on clinicians’ diagnostic confidence and management. Moreover, while a strength of this study was the enrolment of routine patients across three Australian Memory and Cognition Clinics, two-out-of-three clinics were ‘hybrid university/research’ clinics (i.e., clinics embedded within universities, funded via grants and Medicare). These clinics predominantly receive GP referrals from help-seeking individuals for recent onset cognitive and/or mood symptoms, and as such participants enrolled across these sites may not be generalisable to routine patients at Memory and Cognition Clinics that are typically referred directly from clinicians in primary or tertiary care. In addition, these clinics employ a multidisciplinary team for assessment, including a consensus diagnosis meeting, which supports a high level of confidence in initial diagnostic and management decisions. Future studies should thus seek to elucidate the impact of such a blood test on clinician diagnostic confidence and management in settings with solo practitioners. Lastly, among the subsample of Austin Health participants who underwent Aβ-PET imaging revealing a lower than expected Aβ-positivity prevalence, this finding may suggest a bias in participant selection towards those without typical AD presentations, which is supported by the higher overall proportion of participants with SCD and MCI compared to dementia. Practically, a lower disease prevalence improves the negative predicative value while reducing the positive predictive value of diagnostic tests with moderate to high accuracy such as pTau181. Indeed, this low prevalence effect is consistent with the findings from the subsample of participants undergoing Aβ-PET shown in Figure 3. As such, in this study, it may be suggested that plasma pTau181 was more indicative as a diagnostic test to *rule out* rather than *rule in* participants with an underlying AD process, particularly as participants with clinical dementia returned an indeterminant result. Despite this limitation, management impact benefits were demonstrated. Nonetheless, greater clinical benefit is likely to be seen with plasma pTau217, given its superior sensitivity, specificity, and overall diagnostic accuracy for AD.

In conclusion, this study provides novel real-world insights into the clinical implementation of a blood test for AD, demonstrating the impact on clinician decision-making, a key gap which has remained in the literature. Overall, our findings highlight that disclosure of a patient’s AD probability based on plasma pTau181 levels can increase clinician confidence in their diagnosis and management in cases with diagnostic uncertainty (i.e., low-to-moderate confidence). Moreover, while two-thirds of participating clinicians supported the clinical implementation of a blood test for AD, further refinement is needed, particularly around the validation of more sensitive biomarkers such as plasma pTau217. Challenges also remain regarding clinical interpretation, potential costs to the patient, accessibility and infrastructure readiness. While further large-scale studies are needed to build on the methodological framework of this real-world study, our findings mark significant progress toward the clinical implementation of a blood test for AD, paving the way for more accessible and scalable diagnostic approaches in routine care.

## Data Availability

All data produced in the present work are contained in the manuscript.

## ACKNOWLEDGEMENTS

The authors would like to acknowledge Kaylee Rudd for her coordination of the study at the ISLAND clinic; Kirsty Lansley for her assistance with phlebotomy at the Healthy Brain Ageing (HBA) clinic; and Marjorie Garcia, Svetlana Bozinovski, and Beth Veevers for their support with participant recruitment and coordination at the Austin Health Cognitive Dementia and Memory Service (CDAMS).

## CONSENT STATEMENT

All study participants provided informed consent for the collection and use of their data.

